# Efficacy Of Connective Tissue Graft Versus Titanium Papillary Inserts in the Surgical Reconstruction of Interdental Papilla: A Randomized Controlled Clinical Trial

**DOI:** 10.64898/2026.02.26.26345466

**Authors:** Shrushti S Nagar, Rampalli Viswa Chandra, Amarender Reddy Aileni, Vallabhdas Santosh Goud

## Abstract

**Aim and Objectives:** The study aimed to evaluate the effectiveness of titanium inserts for interdental papilla reconstruction, comparing it with the Han and Takei technique using subepithelial connective tissue grafts. The objectives included assessing the black triangle height, papilla height and papilla presence index (PPI) at baseline, 1 month and 3 months postoperatively along with the evaluation of Early Wound Healing Score (EHS) during the first week of post operative healing period.

**Patients and Methods:** This single-blind randomized clinical trial included systemically healthy individuals aged 18–35 years with Nordland and Tarnow’s Class I–III papillary loss. A total of 18 participants were randomly assigned to either test group or control group. Clinical parameters were measured pre- and post-operatively at specified intervals. Both groups received standard presurgical care and postoperative follow-up. The surgical protocol for the test group involved titanium insert placement in the interdental bone, while the control group received a connective tissue graft using the Han and Takei method.

**Results:** Both groups showed significant intragroup improvements in all parameters from baseline to 1 and 3 months (p<0.05). However, intergroup comparisons showed no significant differences at most time points, except at 3 months for PPI, where the control group showed significantly better results (p=0.04). EHS scores were not significant between the groups.

**Conclusion:** Titanium inserts and CTG both demonstrated clinical effectiveness in enhancing interdental papilla dimensions. These findings support the titanium insert as a viable, less invasive alternative, offering clinicians a practical option for esthetic papilla reconstruction.

## INTRODUCTION

The human gingiva consists of specific anatomical components: the attached gingiva, which securely connects the gingiva to the underlying bone, and the marginal (free) gingiva, a slender strip of tissue that surrounds the tooth without adhering to hard tissue. Vertically, the marginal gingiva stretches from the (Cemento-enamel junction) CEJ to the free gingival groove. In the interdental space lies the interdental papilla, a vital part of the gingival structure.^1,2^

Morphologically, the interdental papilla reaches from the interdental alveolar bone to the contact point of neighboring teeth. It is separated from the mesial and distal aspects by sulcular and/or junctional epithelium, whereas the facial and oral surfaces are covered by keratinized gingival oral epithelium. The development of the interdental papilla takes place during tooth eruption, and its shape is affected by the contact points of neighboring teeth, the widths of proximal tooth surfaces, and the configuration of the CEJ.^1,2^

The interdental papilla can be vulnerable to inflammatory agents through the junctional/sulcular epithelia, leading to localized inflammation even in clinically healthy papillae, which is a frequently observed histologic finding.

A black triangle, which denotes the loss of interdental papillae, continues to be one of the most prevalent conditions leading to not just aesthetic and phonetic issues but also functional challenges as it increases the likelihood of food impaction that negatively impacts periodontal health. The interdental papilla in the incisor area is typically pyramidal in form. Elements that affect the presence or absence of interdental papilla include crestal alveolar bone height, the dimensions of the interproximal space, soft tissue appearance (thick or thin biotype), buccal plate thickness, the type of contact area (triangular versus square), and the biologic width.^3^

The limited ability of the interdental papilla to regenerate seems to be influenced by various aspects, such as anatomical factors, bone loss, plaque accumulation, and inflammatory phenomena within the interdental area. Moreover, there is an indication that the interdental papilla may have unique functional features resulting from specific cellular or molecular attributes. This idea is supported by ample documentation showing that distinct anatomical tissue units, even within the marginal gingiva, display specific characteristics. ^4–9^

A range of techniques has been investigated for reconstructing the interdental papilla, encompassing both nonsurgical and surgical strategies. Among the nonsurgical techniques are repeated curettage of the interdental papilla, orthodontic adjustments, restorative fixes, and the application of hyaluronic acid (HA). Although these methods have been tried, they frequently produce less reliable results and provide limited potential for considerable papilla enhancement.

Conversely, surgical approaches have become more prominent due to their effectiveness in papilla reconstruction, despite being more invasive and complex. These surgical options consist of pedicle and free gingival grafts, connective tissue grafts (CTG), subepithelial connective tissue grafts (SCTG), and the application of **Choukroun’s** platelet-rich fibrin (PRF). These techniques involve the manipulation of gum tissues to improve the height and shape of the interdental papilla, targeting more reliable and satisfactory aesthetic results.^10^

Titanium alloys are highly regarded for their outstanding strength-to-weight ratio, resistance to corrosion, and biocompatibility, which makes them suitable for aerospace, medical implants, and marine uses. Their ability to resist oxygen, water, and chemicals guarantees durability in harsh conditions, while their non-toxic nature makes them ideal for medical applications. The characteristics of titanium alloys can be customized through alloying, heat treatment, and surface modifications to satisfy specific application demands.^11^

**El Askary et al** (2000)^12^ described the papillary titanium insert and emphasized its benefits, manipulation, and handling. To support the soft tissue between two implants and achieve the desired interdental papilla-like form, a papillary titanium insert was created. This method eliminates the necessity for any bone grafting procedures while requiring fewer tools and reducing the number of surgical interventions.

This study seeks to present the use of a titanium insert for reconstructing the interdental papilla, which was previously utilized by **El Askary et al**^12^ for forming inter-implant papillae. It also intends to compare this method with the **Han and Takei** technique^13^ of papilla reconstruction using a subepithelial connective tissue graft. The research findings hold the potential to provide clinicians with an innovative therapeutic alternative for enhancing periodontal aesthetics and increasing patient satisfaction.

## MATERIAL AND METHODS

### TRIAL DESIGN

The trial was designed as a single-blind randomized controlled clinical trial to clinically evaluate the use of a titanium insert and subepithelial connective tissue graft for the reconstruction of Interdental papilla.

### PARTICIPANTS AND ELIGIBILITY CRITERIA

Subjects for the study were selected from the outpatient section of the Department of Periodontology, Sri Venkata Sai Institute of Dental Sciences, Mahabubnagar, Telangana and were followed up over three months period after the procedures. The design of the research was approved by Institutional Ethics Committee with approval number SVSIDS/Perio/2/2022. Systemically healthy male and female patients with a stable periodontal health of age 18-35 years. Patients with Nordland and Tarnow’s^13^ Class I, II, III interdental papillary loss at a single site and Gingival recession <2 mm on the facial aspect. Patients presenting with adequate width of attached gingiva (2-3mm) and having a thick gingival biotype (2 mm) were included into the study. Midline diastema patients, medically compromised patients, Pregnant and lactating women, heavy smokers, patient who underwent radiotherapy & chemotherapy, patients with any bleeding or clotting disorders and patients not willing to participate in the study were not included into the study.

### SAMPLE SIZE CALCULATION

Assuming an anticipated prevalence of 36.42%, the necessary sample size is 18 to achieve a margin of error or absolute accuracy of +25% in the prevalence prediction with 95% confidence, taking into account a possible loss or attrition of 15%. At this sample size, the expected 95% CI is 11. 42%. The Scalex SP calculator (Naing L, et. al., 2022) is used to determine this sample size.

### RANDOMIZATION AND BLINDING

In order to assess the effectiveness of "Titanium inserts" in interdental papilla augmentation procedures, the experiment was set up as a randomized controlled clinical trial with single blinding. Randomization included the use of a computer to generate the allocation sequence in random permuted blocks (block randomization), and blinding was achieved by having a second operator assign the block of sites to study groups according to the sequence by which the first operator had coded the two treatment sites selected from each patient into the following groups. Titanium inserts for interdental papilla reconstruction (test group) and connective tissue graft for interdental papilla reconstruction (control group). Until this clinical experiment was over, the blind was not breached.

### INTERVENTIONS

#### Pre-Surgical Protocol

The first phase of treatment for each patient included scaling and root planning, occlusal adjustment, and oral hygiene education in preparation for the procedure. Following the initial examination and thorough phase I treatment, the patients were reevaluated one week later to determine whether they had maintained their oral hygiene. After two weeks, the patients were brought back for the operation.

#### Study Protocol

Before the operation, all baseline parameters (measured on the day of the surgery) were recorded. The following variables were noted: interdental papilla height, Black triangle height, and PPI, which was calculated by **Cardaropoli et al.**^15^, at baseline and again one month and three months after the procedure. The EHS developed by **Marini et al.**^16^ was used one week after surgery to evaluate soft tissue wound healing. All measurements are taken to the closest millimeter.

### SURGICAL PROCEDURE

#### In Test Site

After rinsing with an antiseptic solution for one-minute, adequate amount of local anesthesia was administered. Using a 15C blade, a crevicular incision was made around each tooth, with no incisions across the interdental papilla. Horizontal incision was made at the base of the papilla on the buccal side of the adjacent teeth. Two vertical relaxing incisions were made, parallel to line angle of the adjacent tooth on either side. Semilunar incision was made at the base of the papilla on the palatal side of the interdental space that is to be reconstructed. A full thickness flap was raised apically and the preserved papilla is then elevated with an Orban knife or curettes, intact to the remaining facial flap. After reflection of flap, at the recipient site, a 1-mm diameter twist drill is used to drill midway between the two teeth to a depth of 5 mm. The insert is grasped with pliers and threaded into the osteotomy until it is completely flush with the bone crest which allows for the protrusion of the papilla core of the insert. Soft tissue closure is then achieved through relaxing incisions, and the stripping of the periosteum at the base of the flap allows for a tension-free closure.

#### In Control Site

After rinsing with an antiseptic solution for one-minute, adequate amount of local anesthesia was administered. **Han and Takei** technique^13^ using subepithelial connective tissue graft was planned to be performed. Using a 15-no. blade was used to make a semilunar incision from the disto-labial line angle to the mesio-labial line angle of tooth, at a distance of 6–10 mm in the apical direction from the marginal gingiva in the interdental region. A 12-no. blade was used to make intrasulcular incisions around the mesial half and the distal half of the two adjacent teeth. The gingival-papillary unit was pushed incisally to move gingiva into the crated area to create a split-thickness pouch. The interdental papillary unit was coronally advanced. Subepithelial connective tissue double the size of missing papillary height was harvested from palate. The connective tissue was placed into the pouch space. The semilunar incision was sutured with the interrupted suture, whereas the gingival-papillary unit was sutured coronally using suspensory suture technique.

### POSTSURGICAL CARE

For both the groups, routine postsurgical instructions were given and systemic antibiotics (Amoxicillin-Clavulanate Potassium 625mg thrice daily) was prescribed for 3 days and analgesic (Ibuprofen 800 mg twice daily) for 3 days. A rinse with 10 ml of 0.12% chlorhexidine mouthwash twice daily for 2 weeks was recommended for all the patients. Patients were advised to refrain using any interdental cleansing aids for 4 weeks. Patients were recalled at the end of 1 week for the removal of sutures and every month for oral hygiene maintenance. Clinical measurements were repeated after 1 month and 3-month interval.

### OUTCOMES

#### Primary outcomes

##### Interdental papilla height

It is to be measured from base of the papilla (a line joining the two adjacent teeth gingival zenith) to the tip of the papilla using a UNC-15 probe.

##### Black triangle height

It is to be measured from the tip of the interdental papilla to the apical-most point of the contact point using a UNC-15 probe

##### *Papilla presence index (PPI)* given by Cardaropoli et al: ^15^

The classification system presented here is based on the positional relationship among the papilla, CEJ, and adjacent teeth.

Papilla Presence Index score 1 (PPI 1) is reported when the papilla is completely present and coronally extends to the contact point to completely fill the inter-proximal embrasure. This papilla is at the same level as the adjacent papillae.

Papilla Presence Index score 2 (PPI 2) describes a papilla that is no longer completely present and lies apical to the contact point. This papilla is not at the same level as the adjacent papillae, and the embrasure is no longer completely filled, but the iCEJ is still not visible.

Both PPI 1 and PPI 2 scores can be complicated by the presence of buccal gingival recession, classified as PPI 1r and PPI 2r.

Papilla Presence Index score 3 (PPI 3) refers to the situation in which the papilla is moved more apical and the iCEJ becomes visible. This situation is compatible with a great amount of interdental soft tissue recession.

Papilla Presence Index score 4 (PPI 4) describes when the papilla lies apical to both the iCEJ and buccal CEJ (bCEJ). Interproximal soft tissue recession is present together with buccal gingival recession, and patient esthetics is dramatically compromised.

#### Secondary Outcome

Postoperatively after 1-week, soft tissue wound healing will be assessed by using **Early Wound Healing Score (EHS) by Marini et al**^16^. The EHS scoring was based on the evaluation of clinical signs of re-epithelialization, hemostasis and inflammation. The sum of the single scores for these three parameters calculates the EHS, which ranges between 0 to 10 points.

### STATISTICAL ANALYSIS

The data was analyzed using SPSS© version 22 (IBM, India). Friedman test is used to compare the parameters at various intervals, Post hoc analysis using Bonferroni calculations were used for intra group comparisons and Mann-Whitney U test was used for the inter group comparison of parameters. p value less than 0.05 was set as significant and a value lesser than 0.001 was highly significant.

## OBSERVATIONS AND RESULTS

This was a single-blind randomized clinical trial which included a total of 18 participants who were randomly assigned to either titanium inserts (test group) or connective tissue graft (control group). Clinical parameters were measured pre- and post-operatively at specified intervals that is 1 week, 1 month and 3 months.

### INTRAGROUP COMPARISONS

#### BLACK TRIANGLE HEIGHT

The mean black triangle height *(in mm)* in test group were 2.72 ± 0.75, 3.06± 0.46, 2.00 ± 0.00 and in the control group were 2.17 ± 1.12, 3.44 ± 1.13, 2.00 ± 0.00 at baseline and at the end of 1 and 3 months respectively. (Table 1,2) (Graph 1,2)

**TABLE 1:**
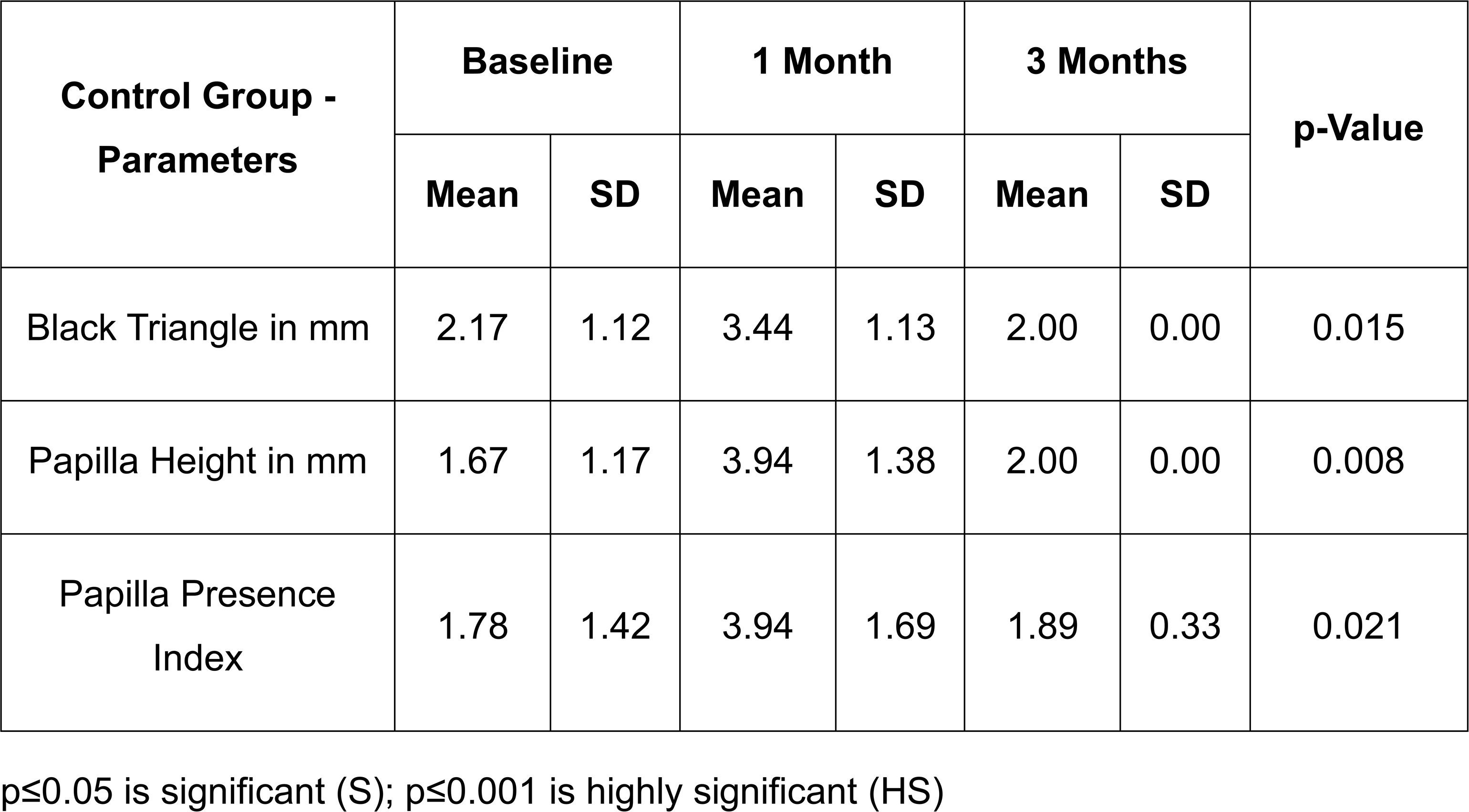
CONTROL GROUP - INTRA GROUP ANALYSIS.

**TABLE 2:**
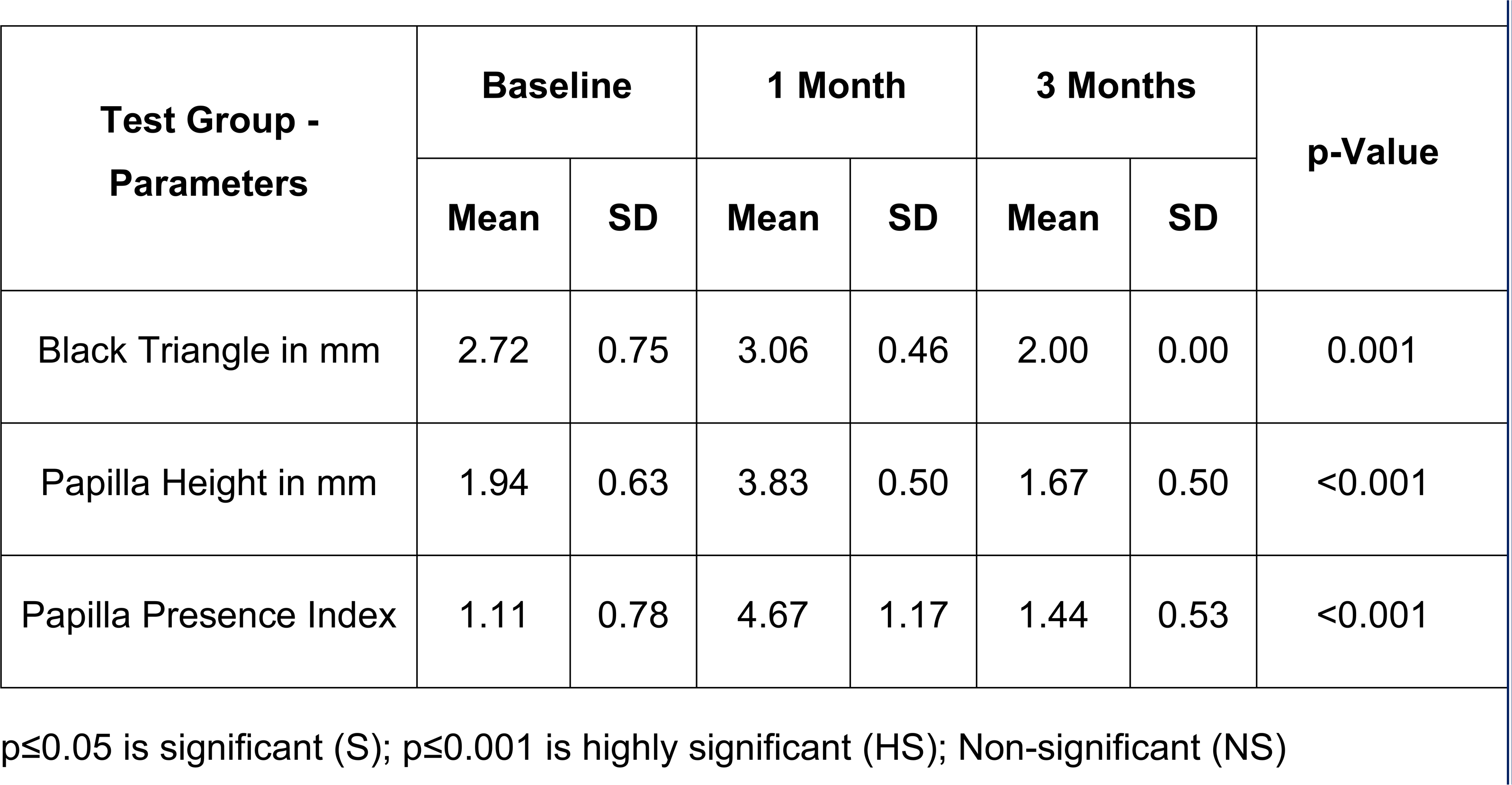
TEST GROUP - INTRA GROUP ANALYSIS.

In both the groups, there was a significant difference seen in the mean black triangle height from the baseline to the end of study period at three-time intervals (p=0.015, p=0.001). (Table 1,2) (Graph 1,2)

#### PAPILLA HEIGHT

The mean papilla height *(in mm)* in test group were 1.94 ± 0.63, 3.83 ± 0.50, 1.67 ± 0.50 and in the control group were 1.67 ± 1.17, 3.94 ± 1.38, 2.00 ± 0.00 at baseline and at the end of 1 and 3 months respectively. (Table 1,2) (Graph 1,2)

There was a significant difference in the mean papilla height in the control group when compared between baseline, 1 month and 3 months (p=0.008). In the test group, there was a highly significant difference seen in the mean papilla height values at all the three time points (p=<0.001). (Table 1,2) (Graph 1,2)

#### PAPILLA PRESENCE INDEX

The mean papilla presence index score in test group were 1.78 ± 1.42, 3.94 ± 1.69, 1.89 ± 0.33 and in the control group were 1.11 ± 0.78, 4.67 ± 1.17, 1.44 ± 0.53 at baseline and at the end of 1 and 3 -months respectively. (Table 1,2) (Graph 1,2)

There was a significant difference in the mean papilla presence index score values in the control group when compared between baseline, 1 month and 3 months. (p=0.021). There was a highly significant difference in the mean papilla presence index scores at three time points in the test group (p=<0.001). (Table 1,2) (Graph 1,2)

### INTERGROUP COMPARISONS

#### BLACK TRIANGLE HEIGHT

There was no significant mean difference between both the groups at baseline, 1 month and 3 months respectively (p=0.23, p=0.54, p=0.23 respectively). (Table 3) (Graph 3)

**TABLE 3:** INTER GROUP ANALYSIS – BLACK TRIANGLE HEIGHT.

| Black Triangle In mm | Test |  | Control |  | p-Value | Significance |
| --- | --- | --- | --- | --- | --- | --- |
|  | Mean | SD | Mean | SD |  |  |
| Baseline | 2.72 | 0.75 | 2.17 | 1.12 | 0.23 | NS |
| 1 Month | 1.94 | 0.63 | 1.67 | 1.17 | 0.54 | NS |
| 3 Months | 1.11 | 0.78 | 1.78 | 1.42 | 0.23 | NS |

$p \leq 0.05$ is significant (S); $p \leq 0.001$ is highly significant (HS); Non-significant (NS)

#### PAPILLA HEIGHT

There was no significant mean difference between both the groups at baseline, 1 month and 3 months respectively (p=0.35, p=0.82, p=0.30 respectively). (Table 4) (Graph 4)

**TABLE 4:** INTER GROUP ANALYSIS – PAPILLARY HEIGHT.

| Papillary Height In mm | Test |  | Control |  | p-Value | Significance |
| --- | --- | --- | --- | --- | --- | --- |
|  | Mean | SD | Mean | SD |  |  |
| Baseline | 3.06 | 0.46 | 3.44 | 1.13 | 0.35 | NS |
| 1 Month | 3.83 | 0.50 | 3.94 | 1.38 | 0.82 | NS |
| 3 Months | 4.67 | 1.17 | 3.94 | 1.69 | 0.30 | NS |

$p \leq 0.05$ is significant (S); $p \leq 0.001$ is highly significant (HS); Non-significant (NS)

#### PAPILLA PRESENCE INDEX

There was no significant mean difference between both the groups at 1 month. (p=0.06). There was a significant mean difference between test and control group, with the mean papillary index of the control group significantly greater than that of the test group (p=0.04). (Table 5) (Graph 5)

**TABLE 5:**
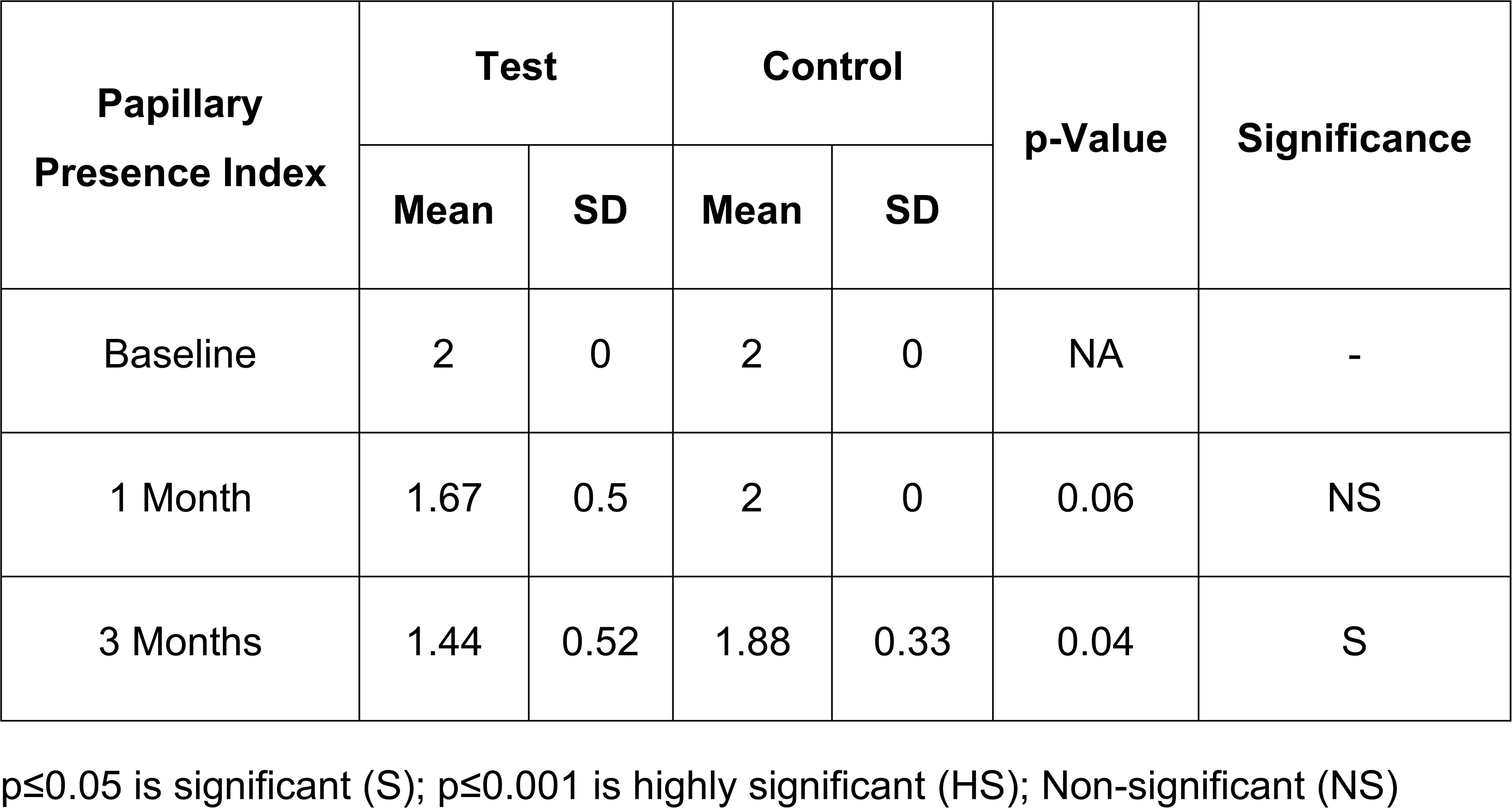
INTER GROUP ANALYSIS – PAPILLA PRESENCE INDEX.

| Papillary<br>Presence Index | Test |  | Control |  | p-Value | Significance |
| --- | --- | --- | --- | --- | --- | --- |
|  | Mean | SD | Mean | SD |  |  |
| Baseline | 2 | 0 | 2 | 0 | NA | - |
| 1 Month | 1.67 | 0.5 | 2 | 0 | 0.06 | NS |
| 3 Months | 1.44 | 0.52 | 1.88 | 0.33 | 0.04 | S |

$p \leq 0.05$ is significant (S); $p \leq 0.001$ is highly significant (HS); Non-significant (NS)

#### EARLY WOUND HEALING SCORE (EHS)

There was no significant mean difference between both the groups at baseline, 1 month and 3 months respectively (p=0.61). (Table 6) (Graph 6)

**TABLE 6:** INTER GROUP ANALYSIS – EARLY WOUND HEALING SCORE.

| Parameter | Test |  | Control |  | P-Value | Significance |
| --- | --- | --- | --- | --- | --- | --- |
|  | Mean | SD | Mean | SD |  |  |
| Early Wound Healing Score | 8.33 | 1.22 | 6.88 | 1.76 | 0.61 | NS |

$p \leq 0.05$ is significant (S); $p \leq 0.001$ is highly significant (HS); Non-significant (NS)

## DISCUSSION

When the interdental papilla is lost due to periodontal disease, surgical interventions, or traumatic incidents, its regenerative capacity is significantly limited compared to that of the marginal or attached gingiva.^17^ Unlike the more predictable healing observed in the marginal regions, where the junctional and sulcular epithelium can effectively re-establish themselves, the interdental papilla presents a unique clinical challenge. Its delicate anatomy and restricted vascularity impede its ability to regenerate fully, thereby making its reconstruction to a form and function that mirrors the original tissue both challenging and unpredictable.^18^

This regenerative discrepancy results in what is known clinically as interdental papillary deficiency or GBT.^16^ These defects not only manifest as noticeable cosmetic deformities that impair the harmony of the smile but also compromise oral function by disrupting speech and encouraging the retention of food debris. Studies have noted that the absence of this critical tissue leaves the periodontium vulnerable to further damage, underscoring the importance of targeted and effective regenerative strategies.^19^

The main objective of periodontal therapy is ‘to prevent the progression of periodontal disease and associated trauma by regeneration of the lost periodontal tissues.^20^ Loss of IDP is multifactorial, and treatment needs a holistic approach concerning the correction of causative factors. Regeneration of IDP, lost from either disease or previous periodontal therapy, is among the most difficult periodontal regenerative treatment, due to its peculiarity of blood supply.^21^

Both non-surgical and surgical approaches have been explored for the management and reconstruction of the interdental papilla. Non-surgical methods^22–23^ aim to correct etiological factors and enhance soft tissue support through conservative means. These include the correction of traumatic oral hygiene practices, restorative modifications to contact points and emergence profiles, orthodontic interventions to optimize interdental spacing, and prosthetic designs such as ovate pontics that promote papillary fill. Additional approaches include repeated curettage to stimulate soft tissue healing and tissue volumizing techniques using dermal fillers or biomaterials to augment deficient papillary areas.

Surgical interventions^22–23^ is often employed when non-surgical methods prove insufficient. These include papilla re-contouring, reconstruction, and preservation. Various preservation techniques have been developed to maintain the integrity of the papilla during surgical procedures. Notable among these are the conventional papilla preservation flap by **Takei et al**. (1985), the modified and simplified versions by **Cortellini et al.** (1995), and the Whale’s Tail technique by **Bianchi and Bassetti** (2009). Reconstruction of the papilla can be achieved using methods such as pedicle flaps, the semilunar coronally repositioned flap introduced by **Tarnow**, envelope-type flaps, autogenous osseous and CTG, and microsurgical techniques, which provide enhanced precision and improved esthetic outcomes.

CTG is a cornerstone procedure in periodontal regenerative therapy, renowned for its predictable outcomes and superior aesthetic and functional results. Originally introduced by **Edel** in the early 1970s, CTG revolutionized soft tissue management by utilizing autogenous tissue—typically harvested from the palatal masticatory mucosa—to promote revascularization, seamless tissue integration, and long-term clinical stability. Its dual blood supply, histologic compatibility, and excellent color match with adjacent tissues have earned it the reputation of the “gold standard” in periodontal plastic surgery.^24^

Despite its many advantages, CTG is not without limitations. The requirement for a secondary donor site increases surgical complexity and may lead to postoperative discomfort, bleeding, or infection. In cases where the palatal vault is shallow, the volume of harvestable tissue may be insufficient for larger reconstructions. Furthermore, the technique-sensitive nature of harvesting and handling CTG necessitates a high level of surgical expertise and careful preoperative planning.^27^ Nevertheless, CTG remains the preferred method for soft tissue augmentation due to its unparalleled histological compatibility, primary healing potential, and long-term clinical success.^24–27^

So, the present *in vivo* randomized clinical study was done to assess the papilla reconstruction in the test group which was compared with the gold standard CTG. The parameters observed for the intragroup analysis were papillary height, black triangle height and papilla presence index. Data obtained by the study were analyzed by using the Friedmans test which was used to compare and check the difference in means in control group with multiple time intervals that is at baseline, 1 month and 3 months. Further post hoc analysis using Bonferroni calculations was done to analyze each parameter and compare it between different time intervals.

The mean black triangle height *(in mm)* in the control group were 2.17 ± 1.12, 3.44 ± 1.13, 2.00 ± 0.00 at baseline and at the end of 1 and 3 months respectively. In control group, there was a significant difference seen in the mean black triangle height from the baseline to the entire study period at the three-time intervals (p=0.015).

The reductions in black triangle height of the current study compares relatively well with a similar study by **Kaushik et al.**^28^, who evaluated the surgical reconstruction of interdental papilla by using an advanced papillary flap with interposed subepithelial connective tissue graft. The mean score of distance from contact point to gingival margin in this study at baseline was 2.60±0.98. At 1st, 3rd and at 6th month the score was 1.87±1.13, with the mean difference of 0.80±0.94 and t-value 3.29 which was statistically significant (p=0.005).

In another study conducted by **Shruthi et al.**^29^, in which the effectiveness of **Han and Takei** technique in reconstructing the lost interdental papilla between maxillary central incisors was checked using papilla height as one of the parameters. The papilla height values decreased from 6.4286 ± 0.97590 at baseline to 4.8571 ± 1.46385 at 6 months which further reduced to 4.5714 ± 1.27242 at 12 months. Statistically significant reduction in PH from baseline was seen. The results of this study match with the present study.

The mean papilla height *(in mm)* in the control group were 1.67 ± 1.17, 3.94 ± 1.38, 2.00 ± 0.00 at baseline and at the end of 1 and 3 months respectively. There was a significant difference in the mean papilla height in the control group when compared between baseline, 1 month and 3 months (p=0.008). Similar results were seen in **Sindhura et al.**^30^ study where it was aimed to evaluate and compare the amount of papillary gain and black triangle height reduction after intervention with a microtunnelling technique with either CTG or PRF as a biomatrix at 6 months using a microsurgical approach. In the intragroup comparison of papillary height showed significant differences in the CTG group.

The mean papilla presence index values in the control group were 1.11 ± 0.78, 4.67 ± 1.17, 1.44 ± 0.53 at baseline and at the end of 1 and 3 months respectively. There was a significant difference in the mean papilla presence index score values in the control group when compared between baseline, 1 month and 3 months (p=0.021). The results coincide with the study done by **Shruthi et al.** ^29^, in which the effectiveness of **Han and Takei** technique in reconstructing the lost interdental papilla between maxillary central incisors was checked using the papilla presence index as one of the parameters. PPI decreased from 2.000 ± 0.000 to 1.5714 ± 0.53452 at 6 months which remained the same at 12 months. There was statistically significant reduction in PPI from baseline.

Titanium is one of the most widely used biomaterials^11^ in medicine and dentistry due to its excellent biocompatibility, corrosion resistance, and favorable mechanical properties. Its ability to form a stable oxide layer makes it highly resistant to corrosion in the human body, while its strength-to-weight ratio and non-toxic nature make it suitable for various applications.^31^ Recent advancements, such as surface modifications and antibacterial coatings, have further enhanced its performance, promoted faster healing and reduced infection risks, ensuring its continued relevance in modern dental practice.^32^

The present study was conducted to evaluate papilla reconstruction using titanium inserts by assessing parameters such as papillary height, black triangle height, and the papilla presence index. The collected data were analyzed using Friedman’s test to compare mean differences within the test group across multiple time intervals—baseline, 1 month, and 3 months. To further assess the significance of changes between each time point, post hoc analysis was performed using Bonferroni correction.

The mean black triangle height *(in mm)* in test group were 2.72 ± 0.75, 3.06± 0.46, 2.00 ± 0.00 at baseline and at the end of 1 and 3 months respectively. There was a significant difference seen in the mean black triangle height from the baseline to the entire study period at the three-time intervals (p=0.001). In another study by **Charde et al.,**^33^ reported that at the 1-year follow-up, complete reconstruction of the interimplant papilla was observed in 90% of the cases. This indicates that using a DFDBA block fixed by a titanium screw is an effective technique for reconstructing the interimplant papilla in the maxillary aesthetic zone during one-stage early loading multiple implant procedures.

The mean papilla height *(in mm)* in test group were 1.94 ± 0.63, 3.83 ± 0.50, 1.67 ± 0.50 at baseline and at the end of 1 and 3 months respectively. There was a highly significant difference in the mean papilla height values at the three time points (p=<0.001). In a case report by **Preeti et al.,**^34^ interimplant papilla reconstruction was performed in the esthetic zone of the maxilla during a one-stage early loading procedure involving multiple implants. The technique utilized a DFDBA block secured with a titanium screw. At baseline, the papillary height measured 2.00 mm. Following the reconstruction, it increased to 3.8 mm at the 3-month mark, showing a gain of 1.8 mm. By the 6 month follow-up, the height further improved to 4.0 mm, resulting in a total gain of 2.0 mm.

The mean papilla presence index score in test group were 1.78 ± 1.42, 3.94 ± 1.69, 1.89 ± 0.33 at baseline and at the end of 1 and 3 months respectively. There was a highly significant difference seen in the mean papilla presence index score values at the three time points (p=<0.001).

Titanium screws offer notable advantages in periodontal regeneration by providing stable and rigid fixation crucial for maintaining space for bone grafts and barrier membranes. Their high modulus of elasticity (generally around 100–110 GPa) minimizes micromovements at the regeneration site, ensuring that the delicate balance of forces during healing is maintained. This rigidity, combined with optimized thread designs, supports secure placement of regenerative materials, thereby aiding in predictable bone fill and soft tissue integration.^35^

Beyond mechanical strength, the inherent biocompatibility of titanium screws is a key asset in periodontal procedures. The naturally occurring titanium oxide layer confers excellent corrosion resistance. Additionally, they exhibit a low affinity for plaque accumulation, which further contributes to a hygienic healing environment. Such properties are critical for the prolonged healing periods typical in guided bone regeneration and other periodontal regenerative approaches.^32^

Keeping in view all these advantages in this study the gold standard CTG was compared with the rigid and biocompatible titanium inserts for papilla augmentation. The use of titanium inserts for papilla augmentation for done previously by **El Askary et al.,**^12^ in inter implant papilla in the anterior maxillary aesthetic region. Here in our study, we are using it for the reconstruction in interdental papilla.

For intergroup analysis, a Man Whitney U test was used was used to compare between the test and control group. There was no significant mean difference in black triangle height between both the groups at baseline, 1 month and 3 months respectively (p=0.23, p=0.54, p=0.23 respectively). There was no significant mean difference in papilla height between both the groups at baseline, 1 month and 3 months respectively (p=0.35, p=0.82, p=0.30 respectively). There was no significant mean difference in papilla presence index between both the groups at 1 month (p=0.06). There was a significant mean difference between test and control group papilla presence index, with the mean papillary index of the control group significantly greater than that of the test group (p=0.04). There was no significant mean difference in the early wound healing score as well between both the groups at baseline, 1 month and 3 months respectively (p=0.61).

Noninferiority trials aim to confirm that a novel treatment performs at least nearly as well as a currently accepted therapy by ensuring that any loss of efficacy does not exceed a clinically acceptable threshold. When the new treatment offers additional benefits, such as increased safety, lower cost, or simpler administration, even if its effectiveness is comparable to the standard therapy, this strategy is especially beneficial. Important considerations in designing these trials include using strict statistical methodologies, establishing a justifiable noninferiority margin based on historical evidence and clinical relevance, and carefully controlling protocol adherence to prevent biases that might wrongly support noninferiority. These considerations are essential to ensuring that the trial results reliably indicate that the new treatment maintains meaningful clinical benefits relative to the established standard.^36^

In this study, the titanium insert was evaluated against CTG, the established gold standard for papillary regeneration. As the comparative analysis revealed no statistically significant differences between the two groups, these findings support the conclusion that this is a non-inferiority study. Therefore, the use of titanium inserts for papilla reconstruction can be considered non-inferior to CTG, demonstrating comparable clinical outcomes without evidence of superiority.

## CONCLUSION AND SUMMARY

A comparative clinical study to evaluate the use of a titanium insert for the reconstruction of Interdental papilla and to compare it with papilla reconstruction using a connective tissue graft. The following conclusions were made with reference to the observation in this study.

1. There is significant difference in all parameters in both the groups when compared between different time intervals.
2. In the present study no statistically significant differences were seen between the two groups concluding the use of titanium inserts for papilla reconstruction can be considered non-inferior to CTG, demonstrating comparable clinical outcomes without evidence of superiority.

**Fig. 1:**
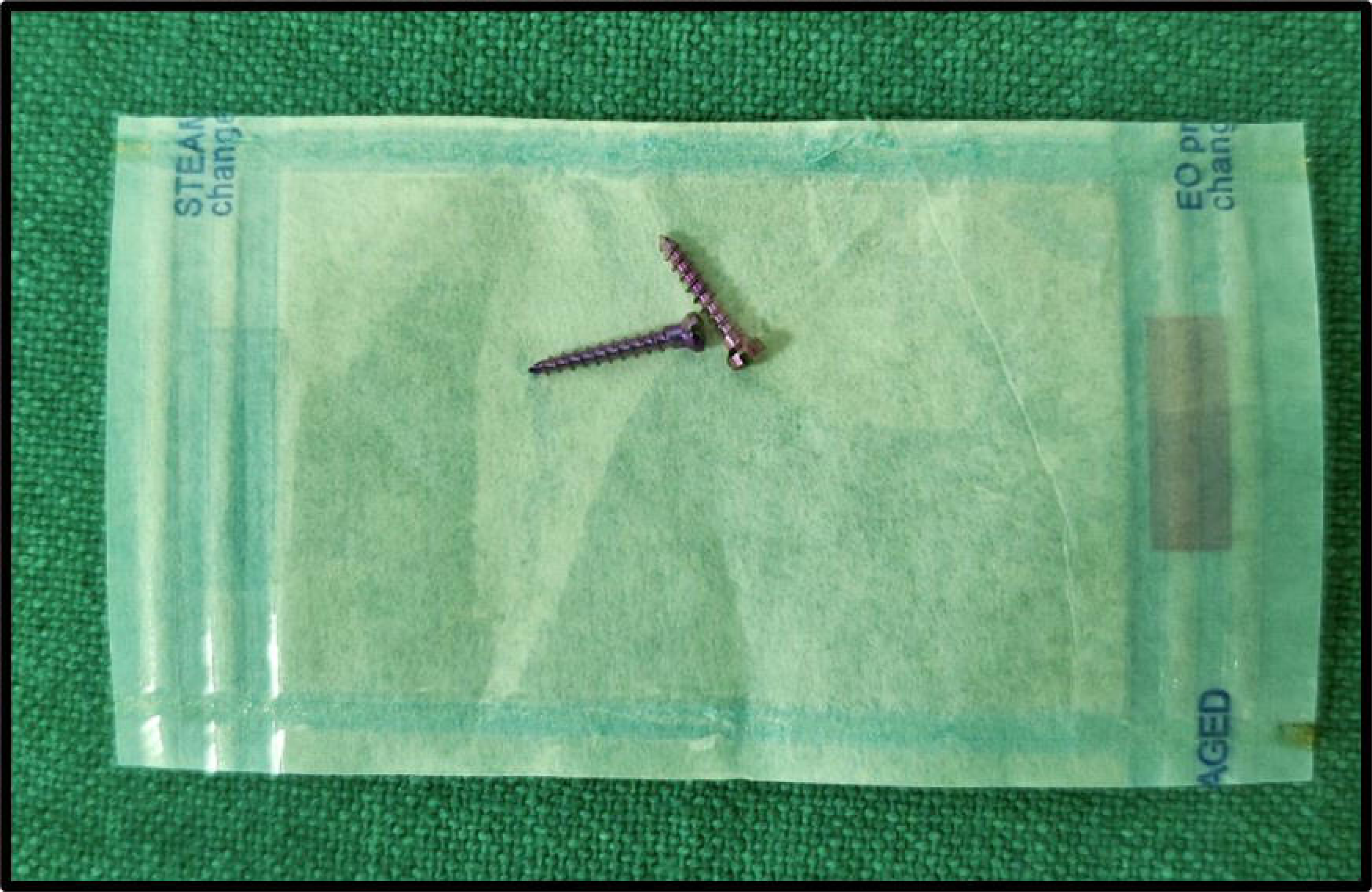
TITANIUM INSERTS

**Fig. 2:**
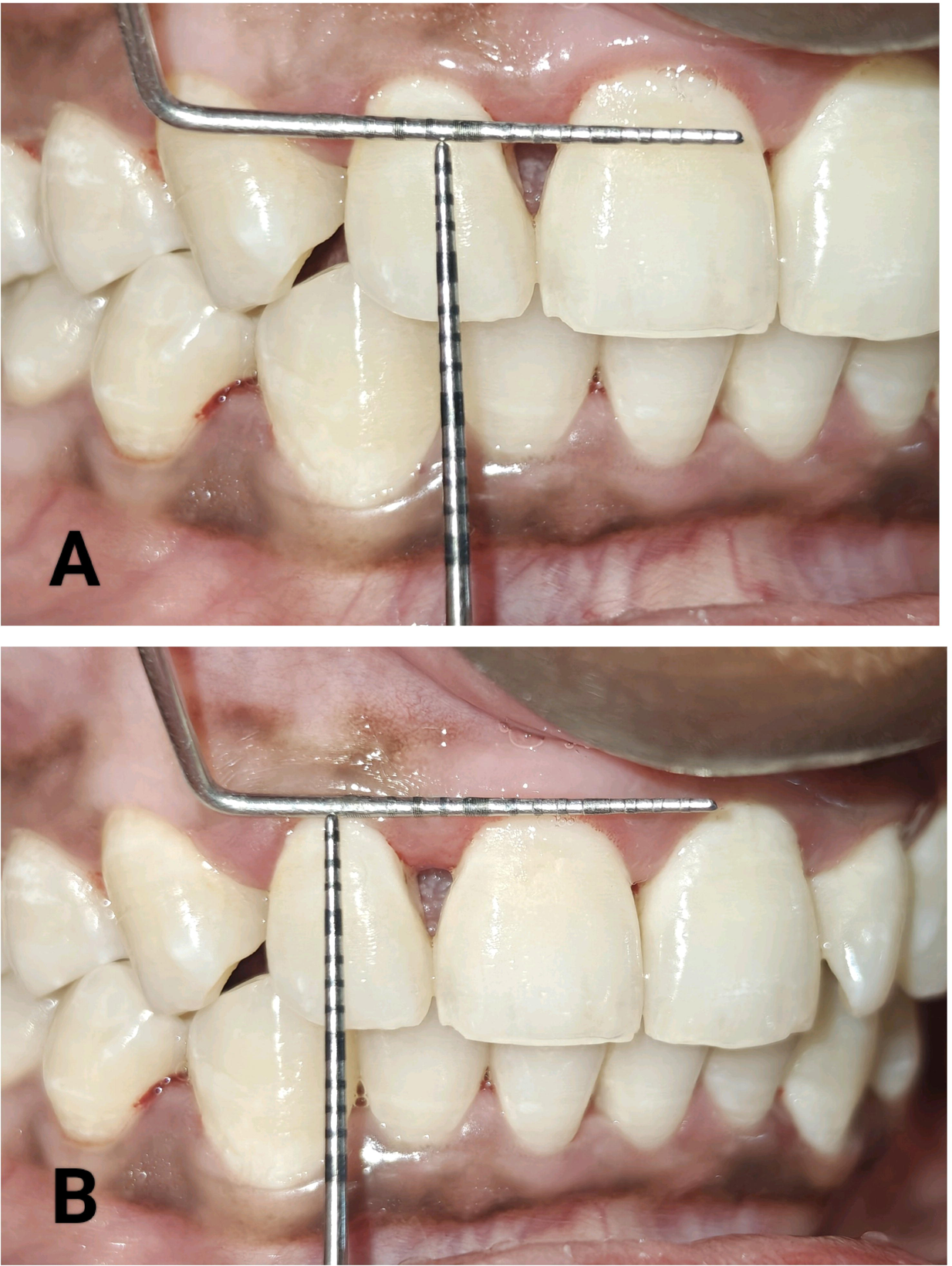
A- PRE – OPERATIVE BLACK TRIANGLE HEIGHT; B- PRE – OPERATIVE BLACK TRIANGLE HEIGHT

**Fig. 3:**
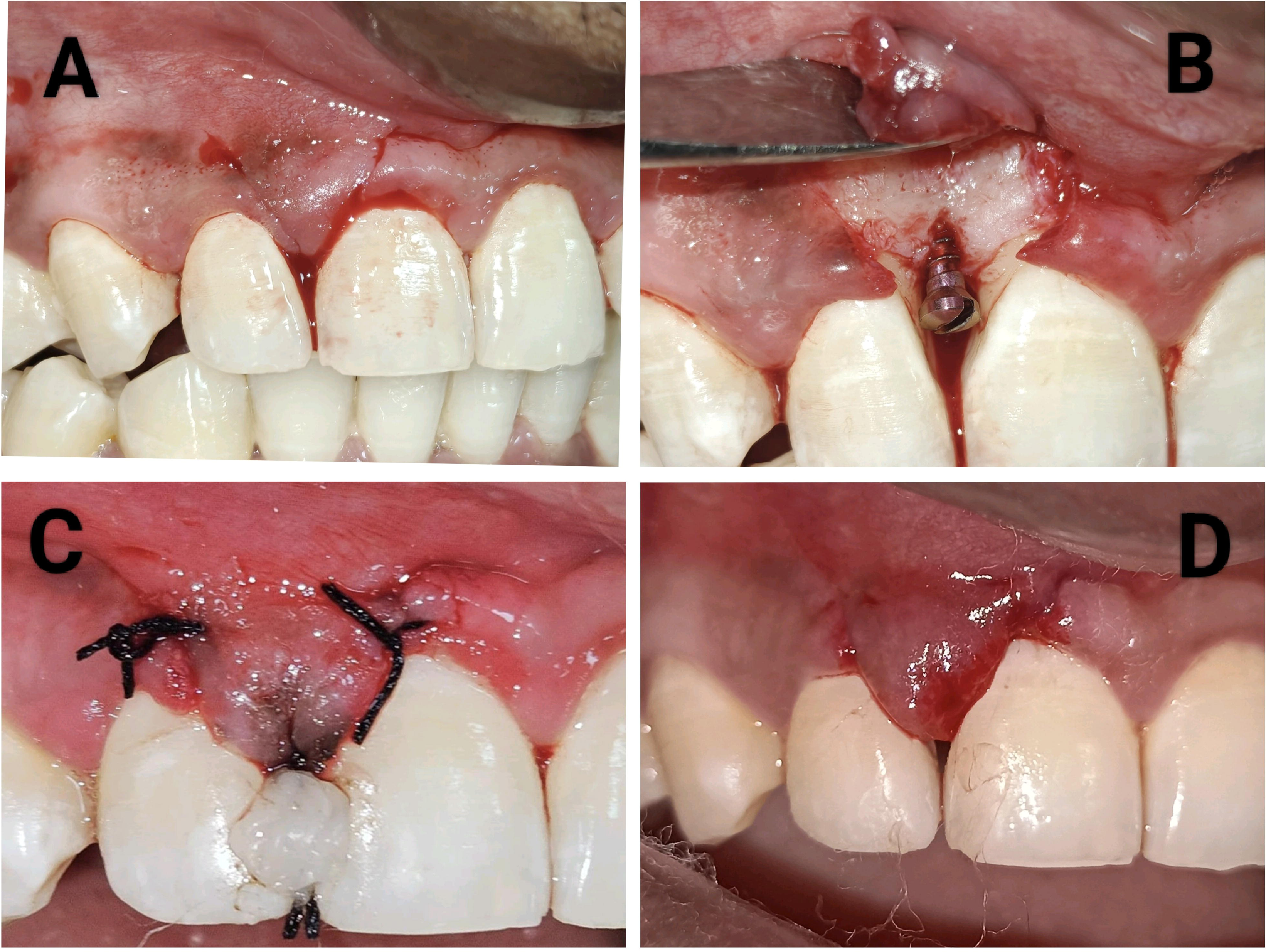
A-INCISION; B- TITANIUM INSERT PLACEMENT; C- SUTURING, D- ONE WEEK POST OPERATIVE

**Fig. 4:**
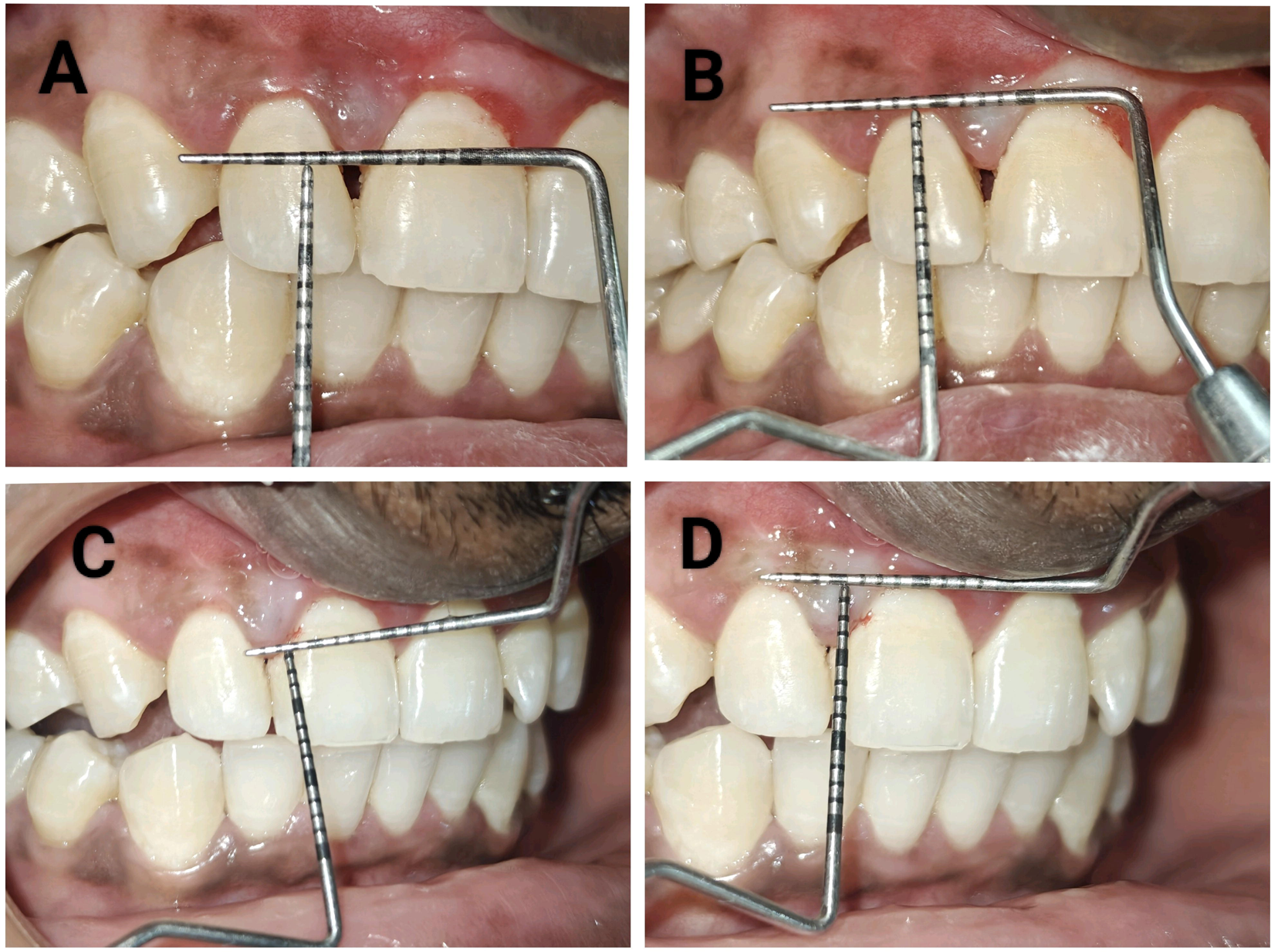
A- 1 MONTH POST – OPERATIVE BLACK TRIANGLE HEIGHT; B- 1 MONTH POST – OPERATIVE PAPILLARY HEIGHT; C- 3 MONTH POST – OPERATIVE BLACK TRAINGLE HEIGHT; D- 3 MONTH POST – OPERATIVE PAPILLARY HEIGHT

**Fig. 5:**
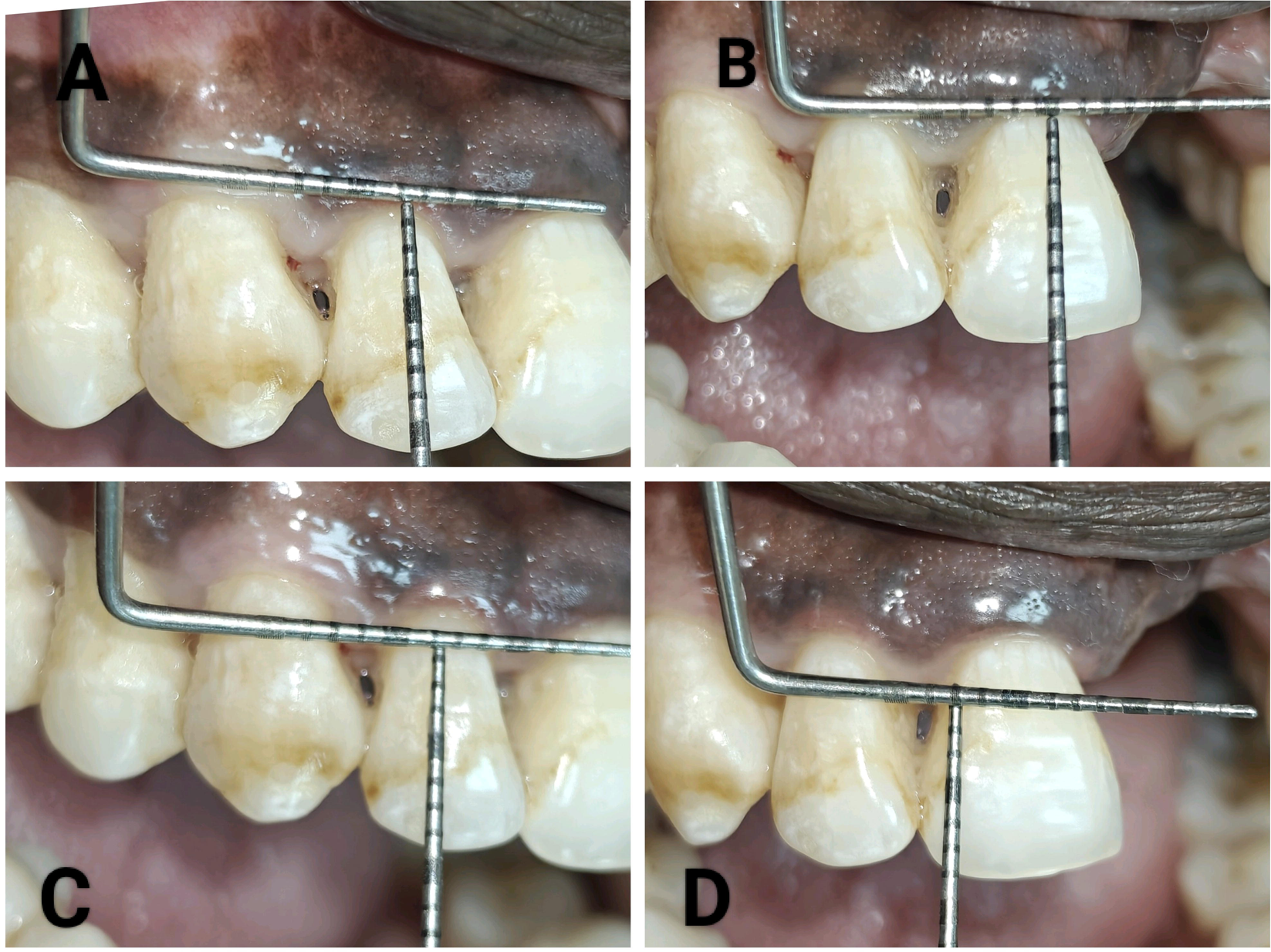
A- PRE – OPERATIVE PAPILLARY HEIGHT; B- PRE – OPERATIVE BLACK TRIANGLE HEIGHT; C-PRE – OPERATIVE PAPILLARY HEIGHT; D- PRE – OPERATIVE BLACK TRIANGLE HEIGHT

**Fig. 6:**
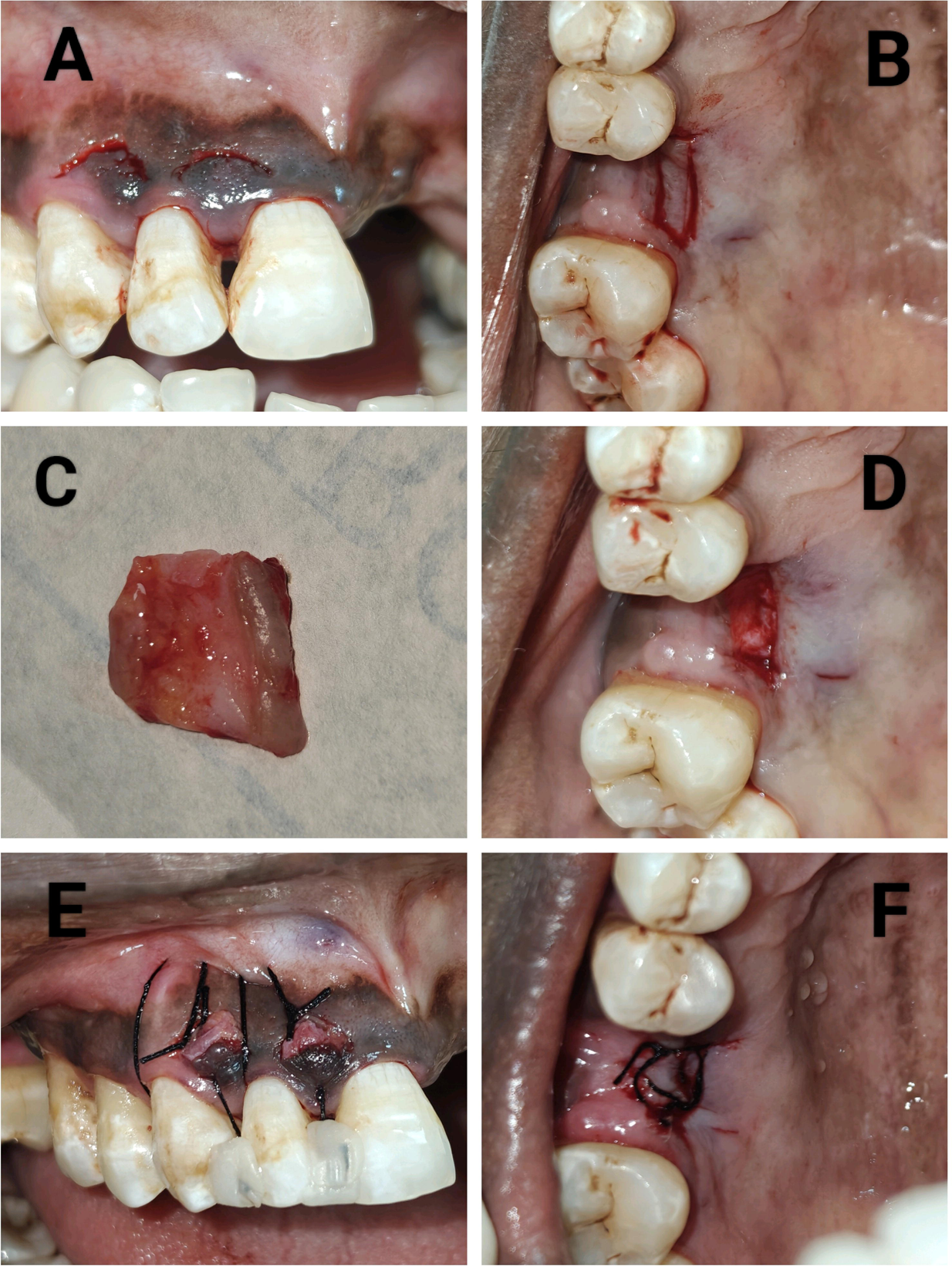
A,B- INCISIONS; C- SCTG HARVESTED; D- DONOR SITE AFTER HARVESTING; E-SUTURING AT RECIPIENT SITE; F- DONOR SITE SUTURING

**Fig. 7:**
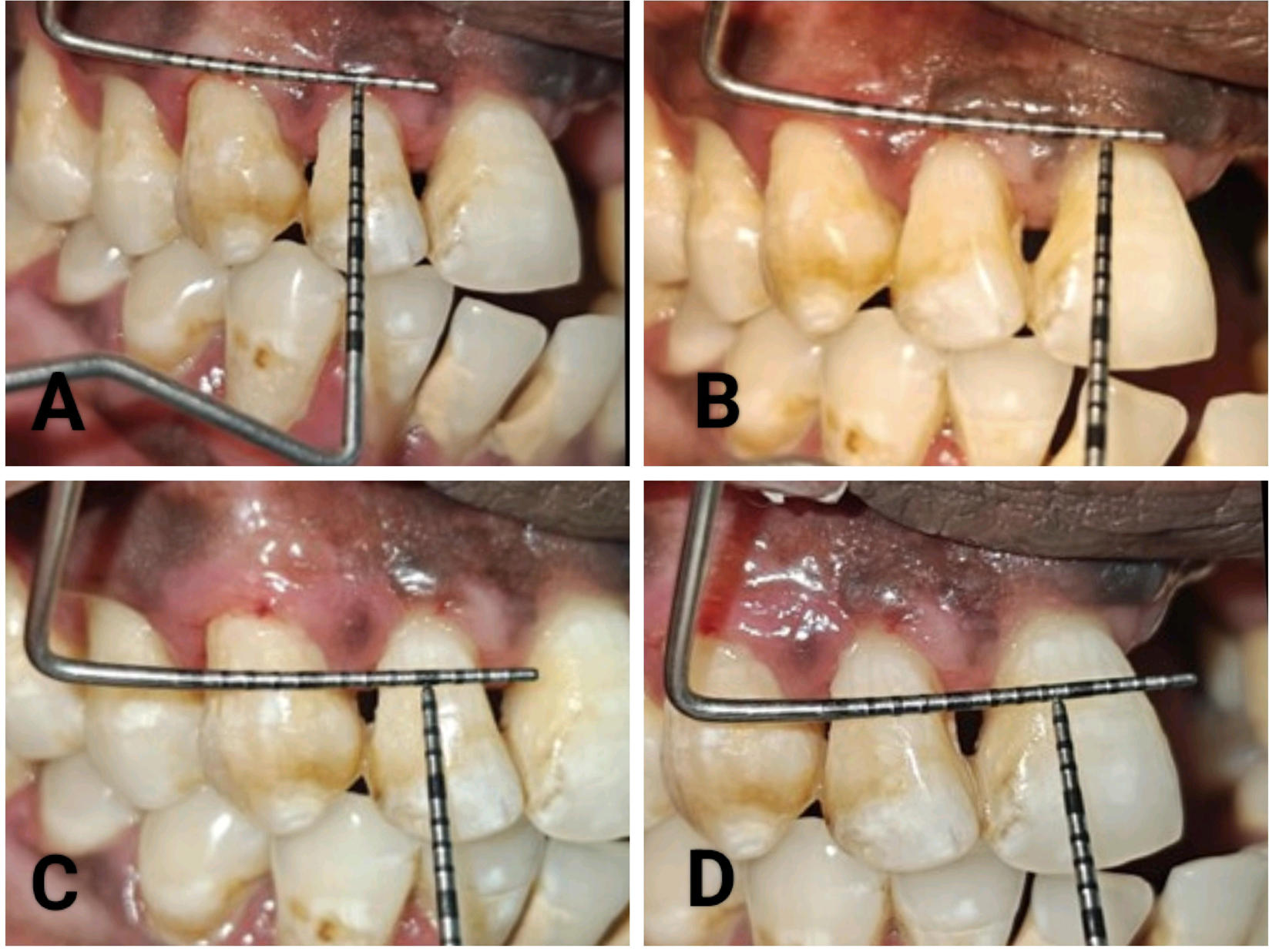
A,B- 1 MONTH POST– OPERATIVE PAPILLARY HEIGHT; C,D- 1 MONTH POST – OPERATIVE BLACK TRIANGLE HEIGHT

**Fig. 8:**
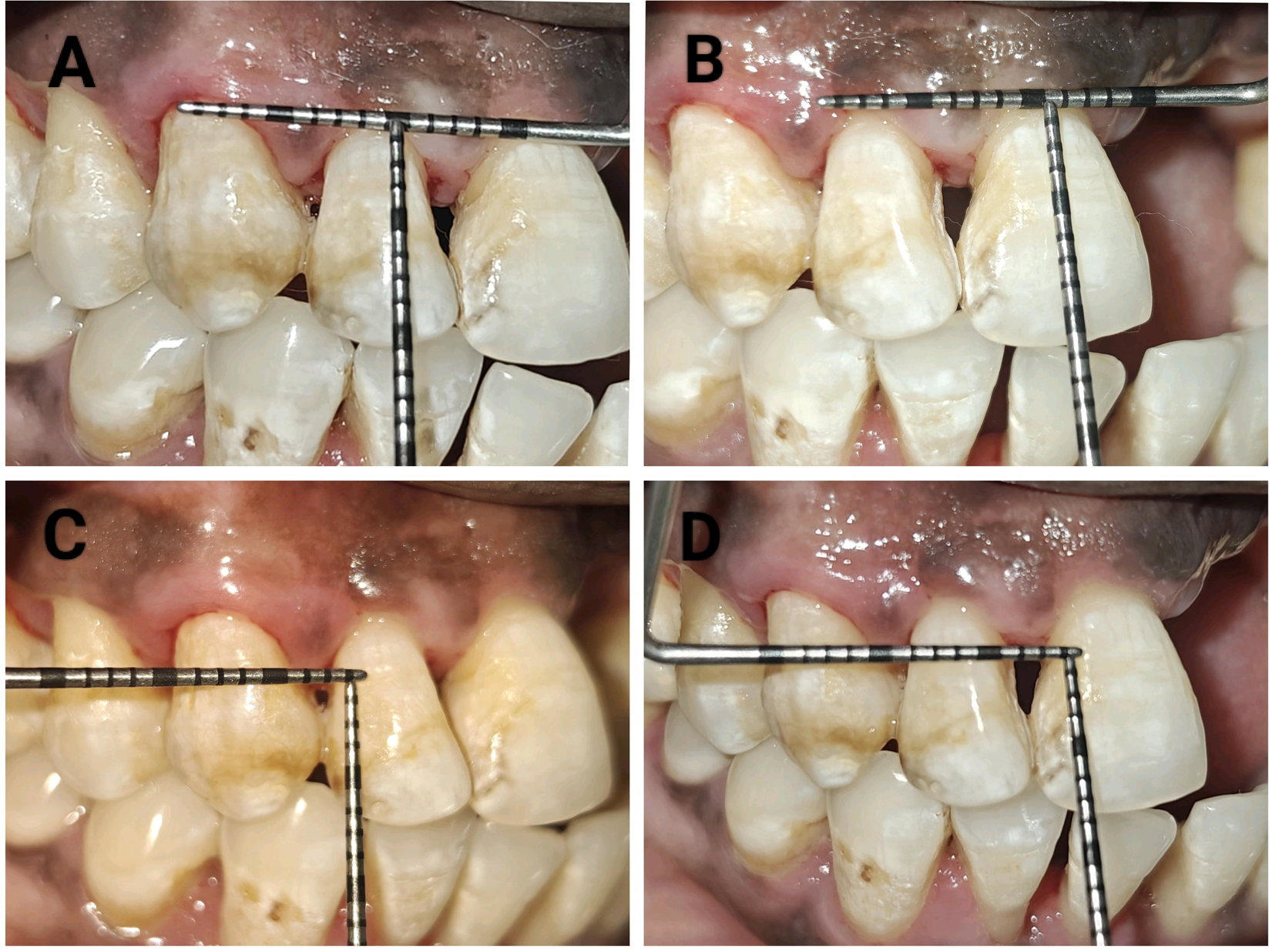
A,B- 3 MONTH POST– OPERATIVE PAPILLARY HEIGHT; C,D- 3 MONTH POST – OPERATIVE BLACK TRIANGLE HEIGHT

## Data Availability

All data produced in the present work are contained in the manuscript

## Notes

### Competing Interest Statement

The authors have declared no competing interest.

### Clinical Trial

NCT07401667

### Funding Statement

This study did not receive any funding

### Author Declarations

Ethics committee of SVS Institute of dental sciences gave ethical approval for this work.

